# SARS-CoV-2 reinfection in a cohort of 43,000 antibody-positive individuals followed for up to 35 weeks

**DOI:** 10.1101/2021.01.15.21249731

**Authors:** Laith J. Abu-Raddad, Hiam Chemaitelly, Peter Coyle, Joel A. Malek, Ayeda A. Ahmed, Yasmin A. Mohamoud, Shameem Younuskunju, Houssein H. Ayoub, Zaina Al Kanaani, Einas Al Kuwari, Adeel A. Butt, Andrew Jeremijenko, Anvar Hassan Kaleeckal, Ali Nizar Latif, Riyazuddin Mohammad Shaik, Hanan F. Abdul Rahim, Gheyath K. Nasrallah, Hadi M. Yassine, Mohamed G. Al Kuwari, Hamad Eid Al Romaihi, Mohamed H. Al-Thani, Abdullatif Al Khal, Roberto Bertollini

**Author notes:** Address reprints requests or correspondence to Professor Laith J. Abu-Raddad, Infectious Disease Epidemiology Group, World Health Organization Collaborating Centre for Disease Epidemiology Analytics on HIV/AIDS, Sexually Transmitted Infections, and Viral Hepatitis, Weill Cornell Medicine - Qatar, Qatar Foundation - Education City, P.O. Box 24144, Doha, Qatar.

## Abstract

**Background:** Reinfection with the severe acute respiratory syndrome coronavirus 2 (SARS-CoV-2) has been documented, raising public health concerns. Risk and incidence rate of SARS-CoV-2 reinfection were assessed in a large cohort of antibody-positive persons in Qatar.

**Methods:** All SARS-CoV-2 antibody-positive persons with a PCR-positive swab ≥14 days after the first-positive antibody test were individually investigated for evidence of reinfection. Viral genome sequencing was conducted for paired viral specimens to confirm reinfection. Incidence of reinfection was compared to incidence of infection in the complement cohort of those antibody-negative.

**Results:** Among 43,044 anti-SARS-CoV-2 positive persons who were followed for a median of 16.3 weeks (range: 0-34.6), 314 individuals (0.7%) had at least one PCR positive swab ≥14 days after the first-positive antibody test. Of these individuals, 129 (41.1%) had supporting epidemiological evidence for reinfection. Reinfection was next investigated using viral genome sequencing. Applying the viral-genome-sequencing confirmation rate, the risk of reinfection was estimated at 0.10% (95% CI: 0.08-0.11%). The incidence rate of reinfection was estimated at 0.66 per 10,000 person-weeks (95% CI: 0.56-0.78). Incidence rate of reinfection versus month of follow-up did not show any evidence of waning of immunity for over seven months of follow-up. Meanwhile, in the complement cohort of 149,923 antibody-negative persons followed for a median of 17.0 weeks (range: 0-45.6), risk of infection was estimated at 2.15% (95% CI: 2.08-2.22%) and incidence rate of infection was estimated at 13.69 per 10,000 person-weeks (95% CI: 13.22-14.14). Efficacy of natural infection against reinfection was estimated at 95.2% (95% CI: 94.1-96.0%). Reinfections were less severe than primary infections. Only one reinfection was severe, two were moderate, and none were critical or fatal. Most reinfections (66.7%) were diagnosed incidentally through random or routine testing, or through contact tracing.

**Conclusions:** Reinfection is rare. Natural infection appears to elicit strong protection against reinfection with an efficacy ∼95% for at least seven months.

## INTRODUCTION

The severe acute respiratory syndrome coronavirus 2 (SARS-CoV-2) pandemic has caused extensive disease and death, with heavy social and economic losses [1-4]. In addition to the risk of first infection, reinfection during this prolonged pandemic has raised additional public health concerns [5-9].

We recently assessed the risk and incidence rate of documented reinfection in a cohort of 130,266 SARS-CoV-2 polymerase chain reaction (PCR)-confirmed infected persons in Qatar [5], a country of 2.8 million population [10, 11] that experienced a large SARS-CoV-2 epidemic [12-16]. Benefiting from a centralized data-capture system for nationwide SARS-CoV-2 PCR testing and using viral genome sequencing, we quantified the risk of reinfection at ∼2 reinfections per 10,000 infected persons [5]. Incidence rate of reinfection was estimated at 0.36 (95% CI: 0.28-0.47) per 10,000 person-weeks [5].

Serological testing for SARS-CoV-2 infection has been expanding in Qatar during the last few months [14, 16, 17]. The first objective of the present study was to quantify the risk and incidence rate of documented reinfection in a cohort of 43,044 persons who had a *laboratory-confirmed, anti-SARS-CoV-2 positive result*, regardless of whether these persons had ever had a diagnosed PCR-confirmed infection. Persons with a *PCR-confirmed* infection could, in principle, be *biologically* different from persons with an *antibody-confirmed* infection, as the former population is more likely to have experienced symptomatic or even serious primary infection, while the latter population is more likely to have experienced an asymptomatic or mild primary infection that may never have been diagnosed. Moreover, some of those with PCR-confirmed infection may not have developed detectable antibodies [5, 7]. The second objective was to estimate the efficacy of natural infection against reinfection by comparing incidence rate of reinfection to incidence rate of infection in the complement cohort of 149,923 persons who had a *laboratory-confirmed, anti-SARS-CoV-2 negative result*.

The present study thus provides an independent assessment of the risk of reinfection in a *biologically* different population from that of PCR-confirmed infected persons. A major strength of the present study is the long follow-up time of each antibody-positive person in this cohort, which had a median of 16.3 weeks for a total cohort follow-up time of 610,832.6 person-weeks, comparable to or greater than the follow-up time in COVID-19 vaccine trials [18-20]. An added strength is the comparison to incidence rate of infection in a large cohort of antibody-negative persons with a similar follow-up time. The study therefore allows assessment of reinfection for more than seven months after primary infection, and provides empirical evidence for possible effects of any waning of immunity.

## METHODS

### Sources of data

We analyzed the centralized and standardized national anti-SARS-CoV-2 serological testing database compiled at Hamad Medical Corporation (HMC), the main public healthcare provider and the nationally designated provider for Coronavirus Disease 2019 (COVID-19) healthcare needs. The database covers essentially all serological testing for SARS-CoV-2 conducted in Qatar, including both testing done on residual blood specimens collected for routine clinical care from attendees at HMC [17] and during a series of population-based serological surveys [14, 16]. The antibody database was linked to the HMC national SARS-CoV-2 PCR testing and COVID-19 hospitalization and severity database [21]. The latter includes records for all SARS-CoV-2 PCR testing conducted in Qatar since the start of the epidemic. The database also includes all COVID-19 hospitalizations and their infection severity classifications, assessed through individual chart reviews by trained medical personnel following World Health Organization (WHO) guidelines [22]. Antibody data were also linked to the centralized COVID-19 death registry, which includes all COVID-19 deaths assessed per WHO guidelines [23].

### Laboratory methods

Antibodies against SARS-CoV-2 in serological samples were detected using the Roche Elecsys^®^ Anti-SARS-CoV-2 assay (Roche, Switzerland), an electrochemiluminescence immunoassay that uses a recombinant protein representing the nucleocapsid (N) antigen for antibody binding. Results were interpreted according to the manufacturer’s instructions (reactive: optical density (proxy for antibody titer [24]) cutoff index ≥1.0 vs. non-reactive: optical density cutoff index <1.0).

Nasopharyngeal and/or oropharyngeal swabs (Huachenyang Technology, China) were collected for PCR testing and placed in Universal Transport Medium (UTM). Aliquots of UTM were: extracted on the QIAsymphony platform (QIAGEN, USA) and tested with real-time reverse-transcription PCR (RT-qPCR) using TaqPath^™^ COVID-19 Combo Kits (Thermo Fisher Scientific, USA) on ABI 7500 FAST (Thermo Fisher, USA). Samples were extracted using a custom protocol [25] on a Hamilton Microlab STAR (Hamilton, USA) and tested using AccuPower SARS-CoV-2 Real-Time RT-PCR Kit (Bioneer, Korea) on ABI 7500 FAST, or loaded directly into a Roche cobas® 6800 system and assayed with a cobas® SARS-CoV-2 Test (Roche, Switzerland). The first assay targets the viral S, N, and ORF1ab regions. The second targets the virus’ RdRp and E-gene regions, and the third targets the ORF1ab and E-gene regions.

All testing was conducted at HMC Central Laboratory or at Sidra Medicine Laboratory, following standardized protocols.

### Suspected reinfection case eligibility and classification

All SARS-CoV-2 antibody-positive persons in Qatar with at least one PCR-positive swab that occurred ≥14 days *after* the first-positive antibody test were considered as *suspected cases* of reinfection. These were classified as showing either *good* evidence, *some* evidence, or *weak* (or *no*) evidence for reinfection based on criteria applied to each case (Box 1). We defined the *reinfection swab* as the first-positive PCR swab that was identified ≥14 days after the first-positive antibody test. The 14-day cutoff was incorporated to exclude cases in which antibody testing and PCR testing were done around the same time as part of clinical care of COVID-19 patients—a PCR-positive swab within few days of an antibody-positive test is likely to reflect active primary infection under clinical consideration rather than a reinfection.

#### Box 1. Classification of suspected cases of SARS-CoV-2 reinfection based on the strength of supporting epidemiological evidence

<u>Suspected cases of SARS-CoV-2 reinfection</u>: All antibody-positive persons with at least one PCR-positive swab that occurred ≥14 days after the first-positive antibody test.

*<u>Good</u>*<u>evidence for reinfection</u>: Individuals who had a PCR-positive swab with a Ct value ≤30 at least 14 days after the first-positive antibody test and who had not had a PCR-positive swab within the 45 days preceding the reinfection swab.

*<u>Some</u>*<u>evidence for reinfection</u>: Individuals who had a PCR-positive swab with a Ct value >30 at least 14 days after the first-positive antibody test and who had not had a PCR-positive swab within the 45 days preceding the reinfection swab.

*<u>Weak</u>*<u>evidence for reinfection</u>: Individuals who had a PCR-positive swab at least 14 days after the first-positive antibody test, but who had one or more PCR-positive swabs within the 45 days preceding the reinfection swab.

Ct, cycle threshold; PCR, polymerase chain reaction.

Suspected reinfection cases with a PCR cycle threshold (Ct) value ≤30 for the reinfection swab (suggestive of a recent active infection) [26-28] and who had *not* had a PCR-positive swab for 45 days preceding the reinfection swab (to rule out persisting PCR positivity due to non-viable virus fragments) [5, 26, 29-31], were considered as showing *good evidence for reinfection*.

Suspected reinfection cases who had not had a PCR-positive swab for 45 days preceding the reinfection swab, but whose Ct value for the reinfection swab was >30, were considered as showing *some evidence for reinfection*.

Suspected reinfection cases who *had* a PCR-positive swab within the 45 days preceding the reinfection swab were considered as showing *weak* (or *no*) evidence for reinfection, as they are likely to reflect prolonged PCR positivity of the primary infection rather than a reinfection [5, 26, 29-31].

### Viral genome sequencing and analysis

For a subset of investigated reinfection cases with *good* or *some evidence for reinfection*, there were records indicating prior diagnosis of the primary infection. Viral genome sequencing was thus conducted to confirm reinfection in this subset of cases whenever it was possible to retrieve both, the first-infection PCR-positive swab and the reinfection swab. Details of viral genome sequencing methods are provided in Supplementary Text S1.

### Reinfection risk and rate

*Risk* of documented reinfection was assessed by quantifying the proportion of cases with *good or some evidence for reinfection* among all eligible anti-SARS-CoV-2 positive cases with an antibody-positive test ≥14 days from end-of-study censoring (excluding cases whose residual blood was tested for antibodies after death).

*Incidence rate* of documented reinfection was calculated by dividing the number of cases with *good* or *some* evidence for reinfection by the number of person-weeks contributed by all anti-SARS-CoV-2 positive cases. The follow-up person-time was calculated starting 14 days after the first-positive antibody test until the reinfection swab, all-cause death, or end-of-study censoring (set on December 31, 2020).

Adjusted estimates for the risk of reinfection and the incidence rate of reinfection were derived by applying the confirmation rate obtained from viral genome sequencing analysis.

### Comparator antibody-negative group and efficacy of natural infection against reinfection

SARS-CoV-2 incidence was also assessed in the complement cohort including all those testing SARS-CoV-2 antibody-negative in Qatar, to provide an antibody-negative comparator group and to assess the efficacy of natural infection against reinfection.

Both *Risk* of documented infection and *Incidence rate* of documented infection in this antibody-negative cohort were assessed as described above for the antibody-positive cohort, but with the *event* defined here as the first PCR-positive swab that is ≥14 days after the first *antibody-negative* test.

The *efficacy* of natural infection against reinfection was estimated by comparing the incidence rate of reinfection in the antibody-positive cohort to the incidence rate of infection in the comparator antibody-negative cohort:

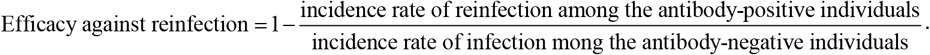

### Ethical approval

This study was approved by the HMC and Weill Cornell Medicine-Qatar Institutional Review Boards.

## RESULTS

### Epidemiological analysis

The process for selecting suspected cases of SARS-CoV-2 reinfection is shown in Figure 1, which summarizes results of their reinfection status evaluation. Of 192,984 persons tested for anti-SARS-CoV-2 using blood specimens collected between April 16-December 31, 2020, 149,934 had negative test results, and were excluded. Six of the remaining 43,050 antibody-positive persons were also excluded because their residual blood was tested for SARS-CoV-2 antibodies after death. This yielded a retrospective cohort of 43,044 antibody-positive persons for whom possible reinfection was assessed.

**Figure 1.**
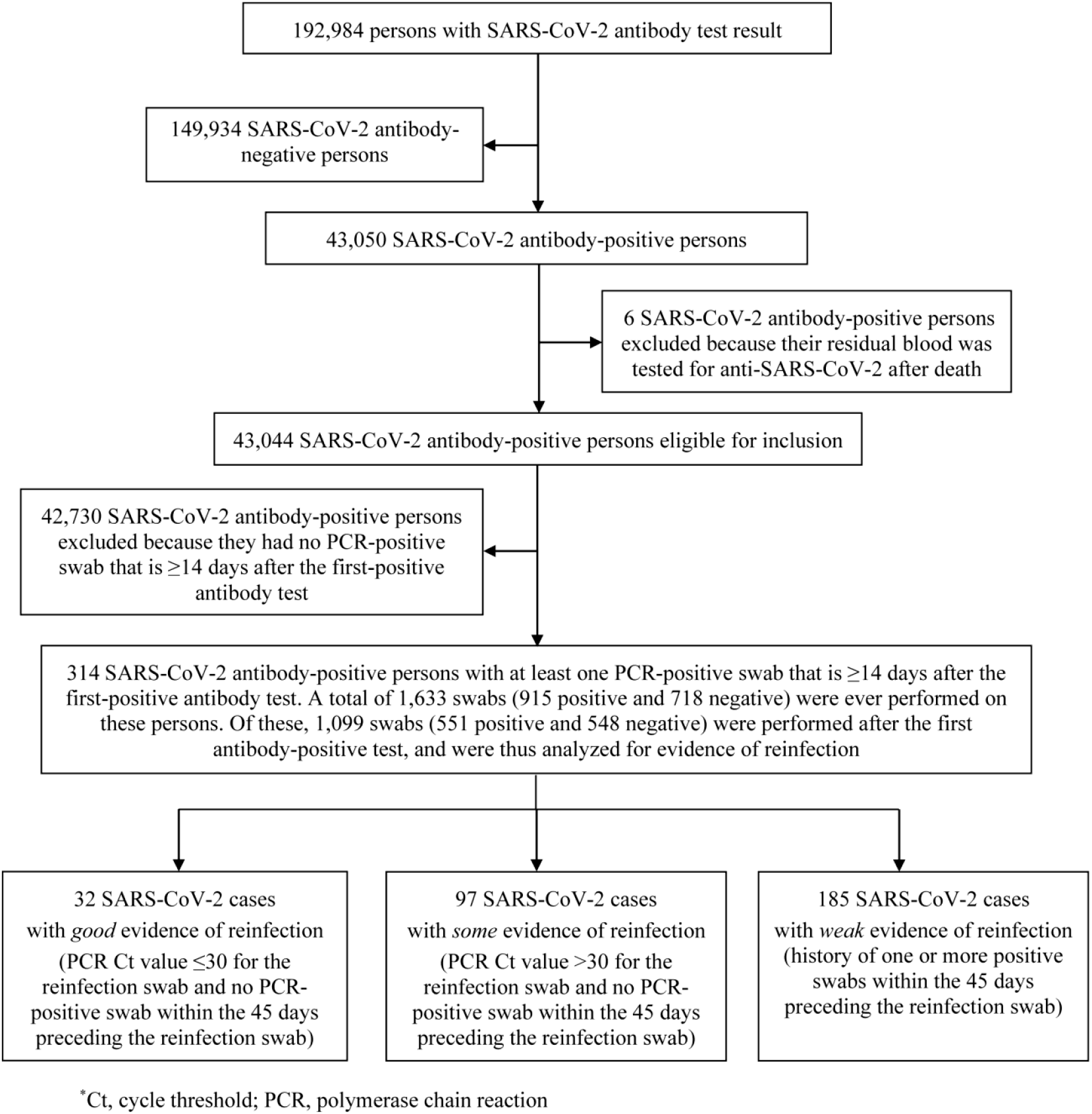
Flow chart describing the selection process of suspected cases of SARS-CoV-2 reinfection and summarizing the results of their reinfection status’ evaluation.

The cohort included 8,953 (20.8%) women and 34,091 men (79.2%) of 158 nationalities. Median age was 35 years for women (interquartile range (IQR): 28-45 years) and 38 years for men (IQR: 31-47 years). Only 19,976 (46.4%) of these persons had ever had a PCR-positive swab *preceding* their first-positive antibody test. Individual time of follow-up ranged between 0 days and 34.6 weeks, with a median of 16.3 weeks.

Only 314 persons had a PCR-positive swab ≥14 days after the first-positive antibody test, and thus qualified for inclusion in the analysis. There were 1,633 swabs (915 positive and 718 negative) collected from these 314 persons, and of these, 1,099 (551 positive and 548 negative) were collected *after* the first-positive antibody test.

Investigation of these 314 suspected cases of reinfection yielded 32 cases with *good* evidence for reinfection (Ct ≤30 for reinfection swab), 97 cases with *some* evidence (Ct >30 for reinfection swab), while evidence was *weak* for the remaining 185 cases.

Characteristics of the 129 cases with *good* or *some* evidence for reinfection are shown in Table 1. These individuals had a median age of 37 years (range: <1-72 years) and included 92 men (71.3%). The median time between the *first-positive antibody* test and the *reinfection* swab was 52 days (range: 15-212 days). The median Ct value of the reinfection swab was 32.9 (range: 13.9-38.3). Slightly over a third of cases were diagnosed based on clinical suspicion (n=34; 26.4%) or individual request (n=9; 7.0%), while the rest (n=86) were identified incidentally either through random PCR-testing campaigns/surveys (n=47; 36.4%), through healthcare routine testing (n=18; 14.0%), through contact tracing (n=15; 11.6%), or at a port of entry (n=6; 4.7%).

**Table 1.**
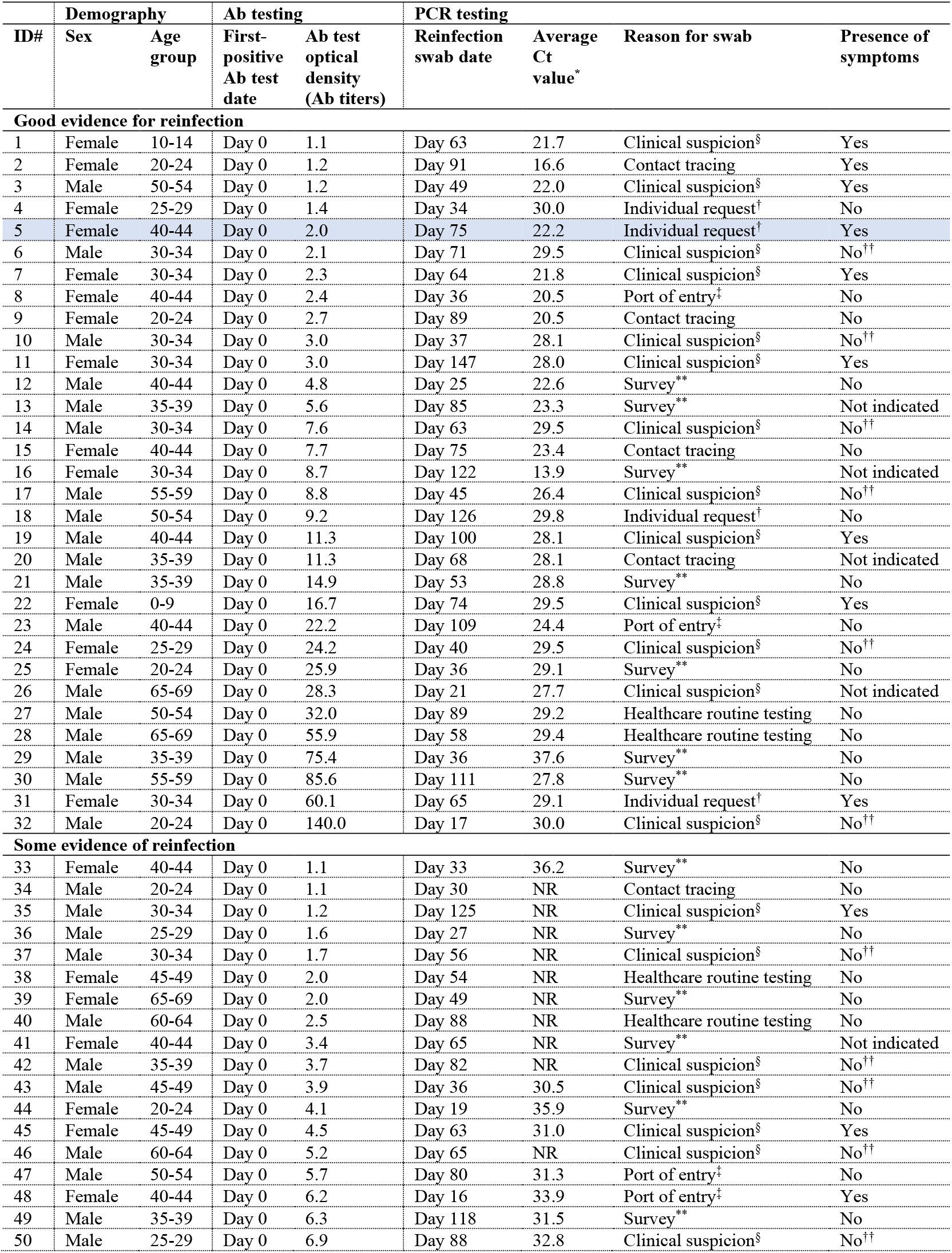
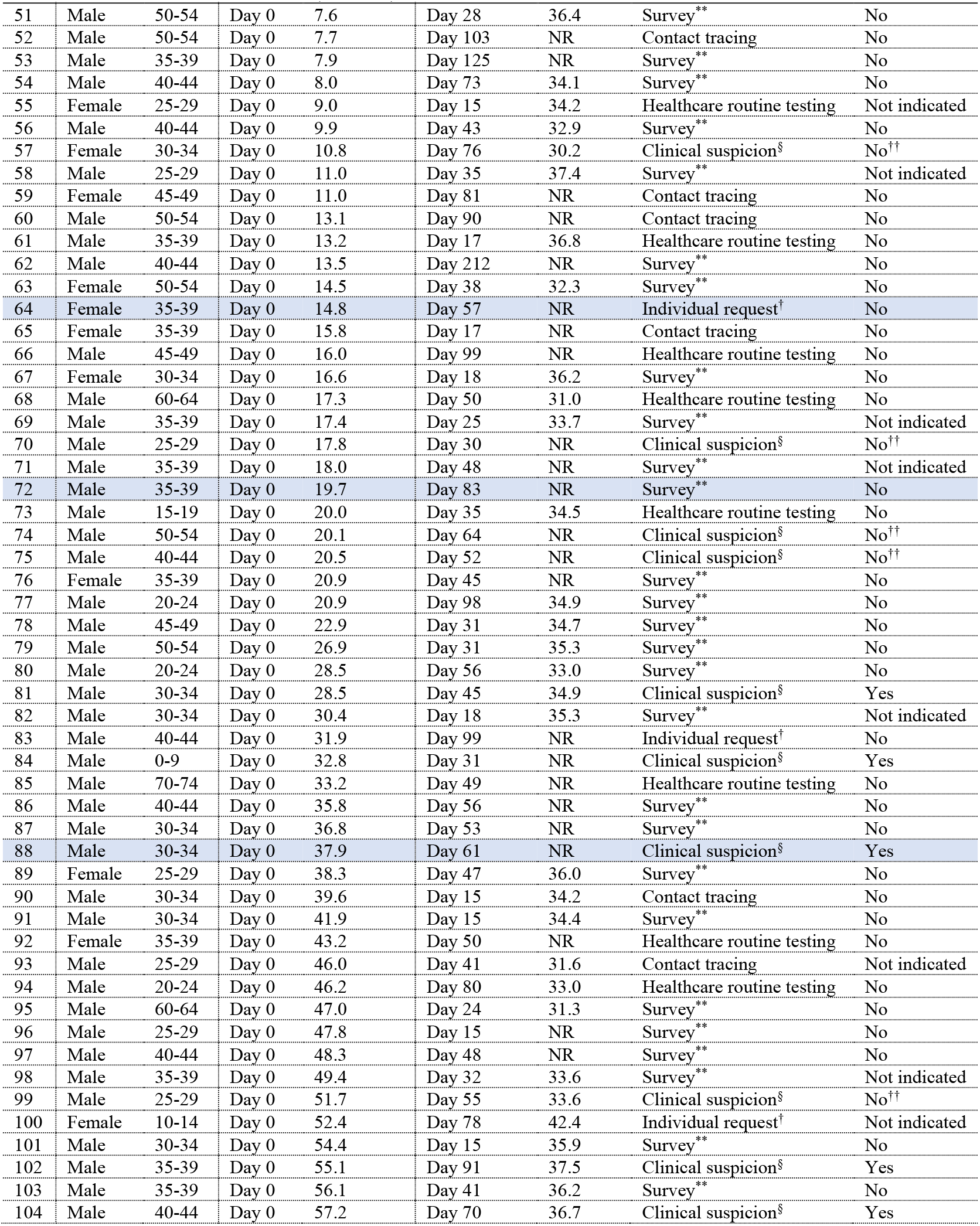
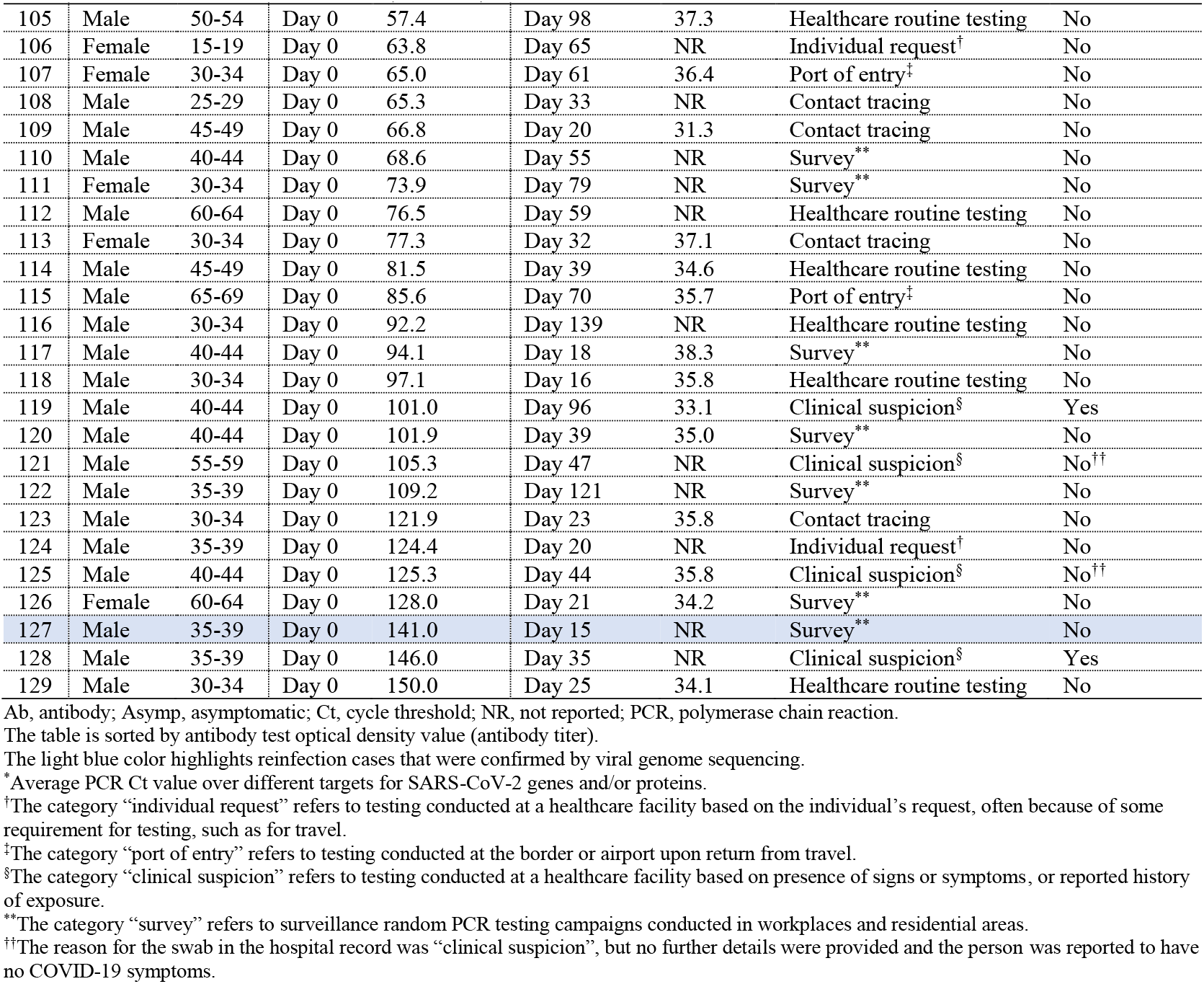
Characteristics of individuals classified as showing good or some evidence of reinfection.

At the time of the reinfection swab, eight cases had records in the severity database. One of these was classified as “severe” and two as “moderate” per WHO classification [22], while the other five were classified as “asymptomatic.” At time of primary infection, 14 cases had records in the severity database, one of whom was classified as “critical”, three as “severe”, five as “moderate”, two as “mild”, and three as “asymptomatic.” For the rest of the reinfection cases, no severity classification was conducted because of minimal or no symptoms to warrant a clinical assessment. For the eight asymptomatic cases above that *had* a severity assessment, the assessment was conducted because of non-COVID-related hospitalization. No deaths were recorded for any of these reinfection cases.

### Confirmation of reinfection through viral genome sequencing

Among the 129 cases with *good* or *some* evidence for reinfection, 62 had records indicating prior diagnosis of a primary infection. Paired specimens of the first-infection PCR-positive swab and the reinfection swab were retrieved in 23 cases. Viral genome sequencing results are summarized in Table 2. Detailed analysis for each genome pair is shown in Figure 2 and Supplementary Figures S1-S2.

**Table 2.** Results of reinfection confirmation using viral genome sequencing. Viral genome sequencing was conducted only for a subset of cases with *good* or *some* evidence of reinfection, that is, whenever paired samples of the first-infection PCR-positive swab and the reinfection PCR-positive swab were available.

| **Viral genome sequencing evidence for reinfection** | **Indication upon comparing each genome pair** | **N** |
| --- | --- | --- |
| Insufficient evidence to warrant interpretation | One or two genomes of low quality | 7 |
| No evidence for reinfection | One change of allele frequency | 1 |
| Shifting balance of quasi-species with no evidence for reinfection | Few changes of allele frequency but not sufficiently indicative of reinfection | 6 |
| Strong evidence for no reinfection | Both genomes of high quality yet no significant differences found | 4* |
| Supporting evidence for reinfection | Few changes of allele frequency indicative of reinfection | 1 |
| Strong evidence for reinfection | Multiple changes of allele frequency indicative of reinfection | 4 |
| **Total** |  | **23** |

PCR, polymerase chain reaction.
\*Viral genome sequencing for two patients was performed as part of an earlier study assessing the risk of SARS-CoV-2 reinfection in the cohort of PCR-confirmed infected persons in Qatar [5].

**Figure 2.**
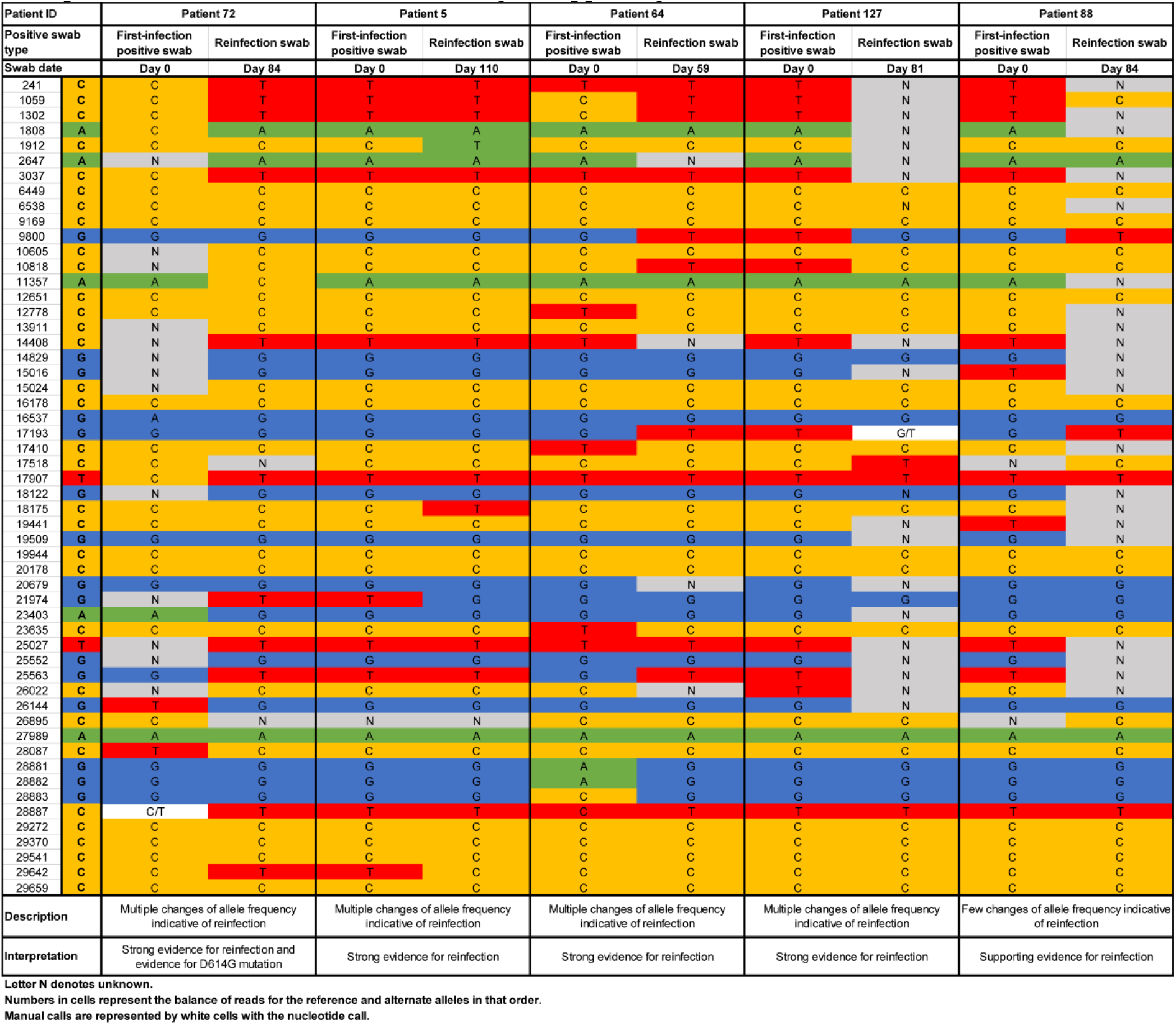
Viral genome sequencing analysis of paired viral specimens of the primary-infection PCR-positive swab and the reinfection PCR-positive swab for five cases with strong or supporting evidence of reinfection.

There was insufficient evidence to warrant interpretation for seven sample pairs because of low genome quality. For seven additional pairs, there were one to several changes of allele frequency indicative at best of a shifting balance of quasi-species, and thus no evidence for reinfection. For four pairs, there was strong evidence for *no reinfection* as both genomes were of high quality, yet no differences were found. Three of these cases had a Ct <30 for the reinfection swab, indicating persistent active infection (Table 1). Two of these cases were reported earlier as part of a case report documenting the existence of prolonged infections [32].

Meanwhile, for one pair, there were few changes of allele frequency offering *supporting evidence for reinfection*. For four other pairs, there were multiple clear changes of allele frequency indicating *strong evidence for reinfection*. One of the latter pairs also documented the presence of the D614G mutation (23403bp A>G) at the reinfection swab—a variant that has progressively replaced the original D614 form [33, 34].

In summary, for the 16 cases where viral genome sequencing evidence was available, five cases were confirmed as reinfections, a confirmation rate of 31.3%. This confirmation rate was similar to that found in our earlier study of reinfection among those with a PCR-confirmed infection at 33.3% [5].

### Assessment of risk and incidence rate of reinfection

Applying the confirmation rate obtained through viral genome sequencing yielded a risk of documented reinfection of 0.10% (95% confidence interval (CI): 0.08-0.11%)—that is 31.3% of 129 reinfections in the cohort of 42,272 anti-SARS-CoV-2 positive persons with an antibody-positive test ≥14 days from end-of-study censoring.

The incidence rate of documented reinfection was estimated at 0.66 per 10,000 person-weeks (95% CI: 0.56-0.78). That is 31.3% of 129 reinfection events in a follow-up person-time of 610,832.5 person-weeks.

Figure 3 shows the incidence rate of documented reinfection versus month of follow-up in this cohort of antibody-positive persons. There was evidence for a decreasing trend in the incidence rate of reinfection with each additional month of follow-up (Mantel-Haenszel trend analysis p-value: <0.001).

**Figure 3.**
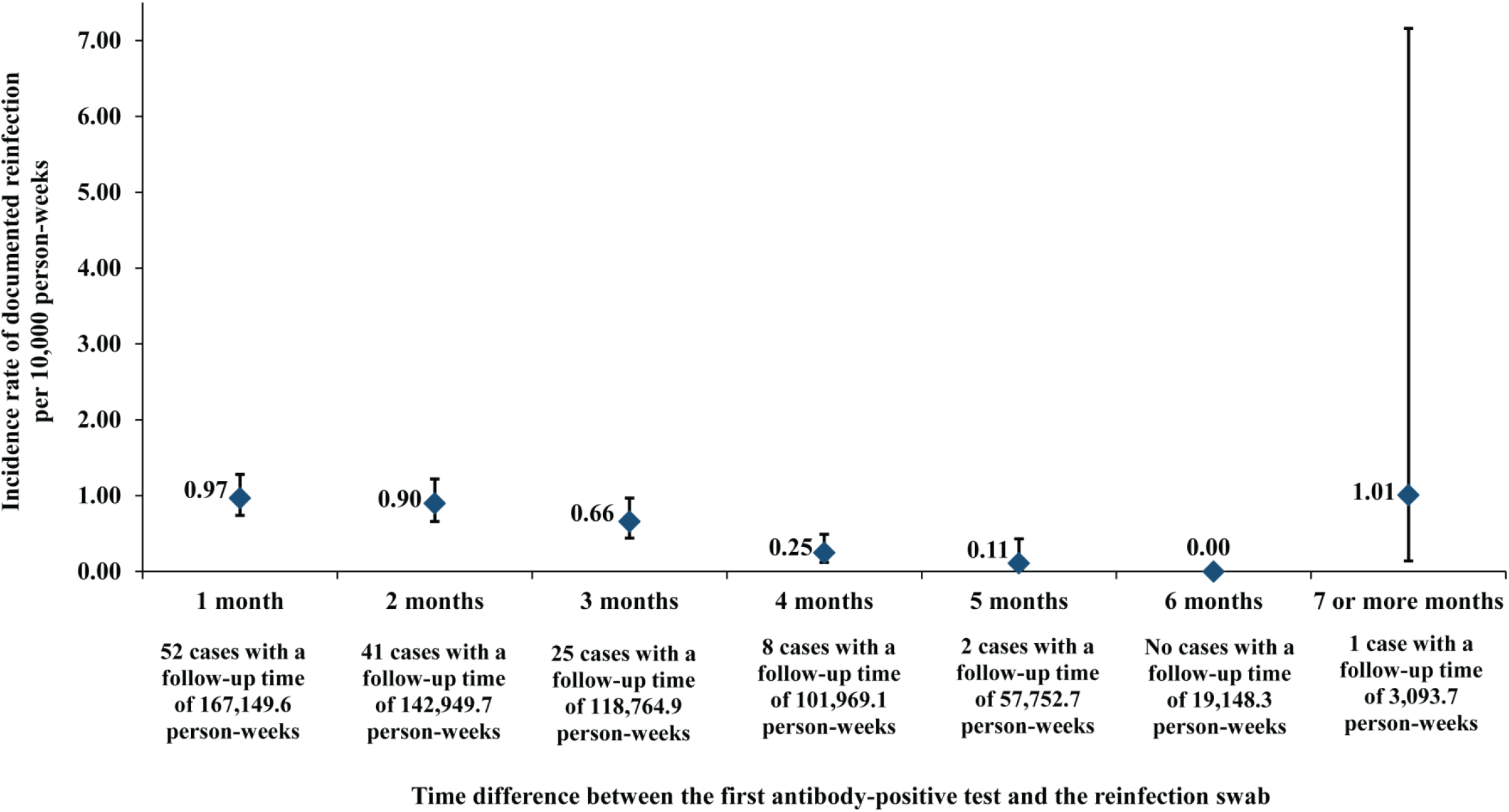
Incidence rate of documented SARS-CoV-2 reinfection versus month of follow-up in the cohort of 43,044 antibody-positive persons.

### Comparator antibody-negative group and efficacy of natural infection against reinfection

The complement cohort of all those testing SARS-CoV-2 antibody-negative included 149,934 individuals. Of those, nine were excluded because their residual blood was tested for SARS-CoV-2 antibodies after death. Two other individuals were excluded because their date of death could not be precisely ascertained. This yielded a retrospective cohort of 149,923 antibody-negative persons to be assessed for SARS-CoV-2 infection incidence (Figure 4).

**Figure 4.**
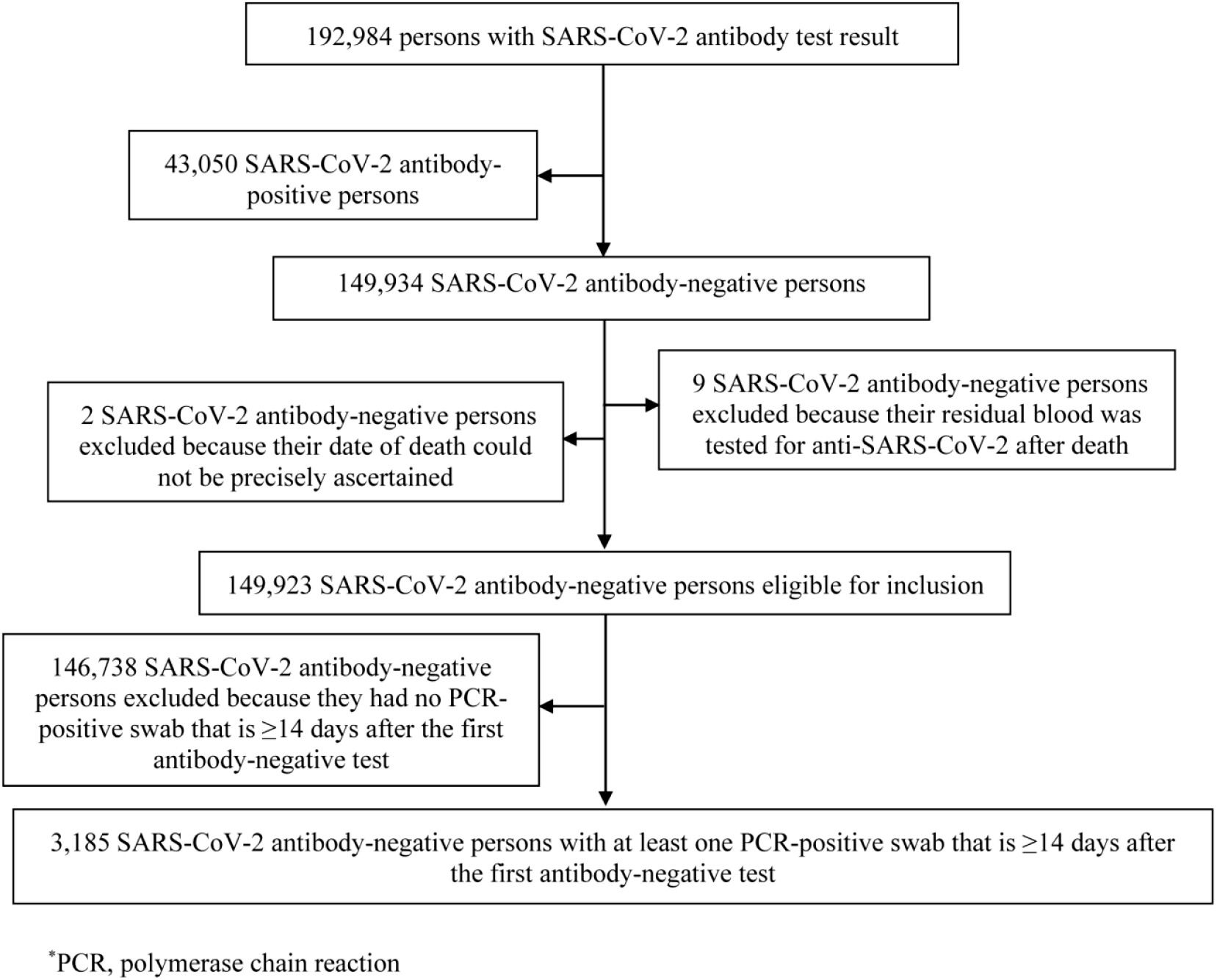
Flow chart describing the process for identifying SARS-CoV-2 incident infections in the complement cohort of antibody-negative individuals.

This cohort included 75,904 (50.6%) women and 74,019 men (49.4%) of 167 nationalities. Median age was 35 years for women (interquartile range (IQR): 28-47 years) and 39 years for men (IQR: 30-50 years). Individual time of follow-up ranged between 0 days and 45.6 weeks, with a median of 17.0 weeks. These characteristics are similar to those of the antibody-positive cohort apart from the higher proportion of women. The higher proportion of women is a consequence of the fact that men were several-fold more affected than women by the SARS-CoV-2 epidemic in Qatar [12, 14, 16, 17], as the men craft and manual worker population, who comprise 60% of the total population [35], was the most affected with a seroprevalence that is several-fold higher than the rest of the population [12, 14, 16, 17].

Of the 149,923 antibody-negative individuals, 3,185 individuals had at least one PCR-positive swab ≥14 days after the first antibody-negative test. Consequently, the risk of documented infection was estimated at 2.15% (95% CI: 2.08-2.22%)—that is 3,185 infections in the cohort of 148,181 anti-SARS-CoV-2 negative persons with an antibody-negative test ≥14 days from end-of-study censoring.

The incidence rate of documented infection was estimated at 13.69 per 10,000 person-weeks (95% CI: 13.22-14.14), that is 3,185 infections in a follow-up person-time of 2,326,572.0 person-weeks.

The *efficacy* of natural infection against reinfection was estimated by comparing the incidence rate of reinfection in the antibody-positive cohort to the incidence rate of infection in the comparator antibody-negative cohort:

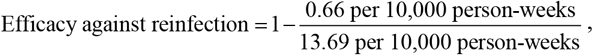

yielding an efficacy estimate of 95.2% (95% CI: 94.1-96.0%).

## DISCUSSION

The results provide concrete evidence for the presence of reinfection in some individuals with detectable antibodies for SARS-CoV-2 infection, even in some with high antibody titers (Table 1). However, the risk of documented reinfection was rare, at ∼1 per 1,000 infected persons, at least for a few months after the first antibody-positive test. There was also no evidence that antibody-positive persons experienced any waning of protective immunity over time, as the incidence rate of reinfection versus month of follow-up did not show an increasing trend over seven months following the first antibody-positive test (Figure 3). To the contrary, there was a trend of decreasing incidence rate, possibly explained by the (very) slowly declining incidence rate in the wider population of Qatar over the last six months [15, 36], or possibly by strengthening of protective immunity due to repeated exposures that did not lead to established infection. Notably, a recent study from Qatar indicated an association between higher antibody titers and repeated exposures to the virus [17]. Further follow up of this cohort of antibody-positive persons over time may allow a more long-term assessment of the persistence of protection against reinfection.

Remarkably, the incidence rate of reinfection found here for those with *antibody-confirmed* infection at ∼1 per 10,000 person-weeks is very similar to that found for those with *PCR-confirmed* infection, as reported in our earlier reinfection study [5]. This suggests that these two populations are *functionally* similar. Evidence of exposure to SARS-CoV-2, regardless of the biomarker used to assess infection, appears sufficient to indicate protection against reinfection.

These findings are striking, as the epidemic in Qatar has been intense, with half of the population estimated to have acquired this infection at some point since its introduction into Qatar early in 2020 [14-17, 36]. It is highly probable that a proportion of the population has been repeatedly exposed to SARS-CoV-2, but such re-exposures did not lead to more than a limited number of documentable reinfections. Other lines of evidence also support a low frequency of reinfection. The epidemic in Qatar grew rapidly and declined rapidly [15, 36], consistent with a susceptible-infected-recovered “SIR” epidemic dynamic in which infection elicits strong immunity against reinfection. No second wave has materialized since the epidemic peaked in May of 2020, despite easing of most restrictions [15, 36]. A recent study on health care workers in the United Kingdom also indicated lower incidence of infection in those antibody-positive [37], and a study of immunological memory in a cohort of COVID-19 patients indicated durability of the immune response for at least 6-8 months [38].

The study estimated the *efficacy* of *natural infection* against *reinfection* at 95.2% by comparing SARS-CoV-2 incidence in those antibody-positive to those antibody-negative. The efficacy can also be estimated by comparing the incidence rate of documented reinfection to the incidence rate of documented infection throughout the epidemic in the total population of Qatar that was estimated at ∼15 per 10,000 person-weeks [15]. This yielded

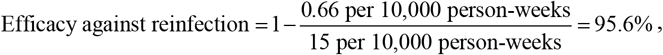

confirming the same efficacy estimate. Remarkably, this efficacy estimate is similar to the efficacy reported for two recently-developed COVID-19 vaccines [18, 19].

While one reinfection was severe, none were critical or fatal and a large proportion of reinfections were minimally symptomatic (if not asymptomatic) to the extent that they were discovered only incidentally, such as through contact tracing or random testing campaigns/surveys (Table 1). The severity of reinfection was also less than that of primary infection. These findings suggest that reinfections (when they rarely occur) appear well tolerated and no more symptomatic than primary infections.

This study has some limitations. By study design, primary infection was *indirectly* ascertained through serological testing, thereby including only a subset with documented PCR-confirmed primary infections. Having said so, serological testing was based on a high-quality, validated platform, the Roche platform, one of the best available and most extensively used and investigated commercial platforms, with a specificity of at least 99.8% [39, 40]. Thus, it is unlikely that misclassified antibody-positives could have biased our findings. The antibody-negative cohort had a higher proportion of women than the antibody-positive cohort, due to the differential spread of the infection among women versus men in Qatar [12, 14, 16, 17]. Viral genome sequencing analysis was possible for only a subset of reinfections, either because primary infection was only identified through antibody testing with no record of earlier PCR testing, or because the reinfection swab could not be retrieved. Reinfections were confirmed by noting differences in the viral genome between the primary infection and the reinfection. While not likely, it is theoretically possible that these differences may have occurred due to within-host evolution of the virus, as in the context of a prolonged infection [32, 41]. The potential effect of these limitations is likely an overestimation, rather than underestimation, of the incidence of reinfection, thereby affirming the conclusion of the rarity of reinfections.

In conclusion, SARS-CoV-2 reinfection was investigated in a large cohort of antibody-positive individuals who were followed for as long as 35 weeks. While the study documented some reinfections, they constitute a rare phenomenon, with natural infection eliciting protection against reinfection with an efficacy of ∼95%. This points to development of robust immunity following primary infection, which lasts for at least seven months. These findings may suggest that prioritizing vaccination for those who are antibody-negative, as long as doses of the vaccine remain in short supply, could enhance the health, societal, and economic gains attained by vaccination.

## Data Availability

All data are available within the manuscript and supplementary materials.

## Acknowledgements

We thank Her Excellency Dr. Hanan Al Kuwari, Minister of Public Health, for her vision, guidance, leadership, and support. We also thank Dr. Saad Al Kaabi, Chair of the System Wide Incident Command and Control (SWICC) Committee for the COVID-19 national healthcare response, for his leadership, analytical insights, and for his instrumental role in enacting data information systems that made these studies possible. We further extend our appreciation to the SWICC Committee and the Scientific Reference and Research Taskforce (SRRT) members for their informative input, scientific technical advice, and enriching discussions. We also thank Dr. Mariam Abdulmalik, CEO of the Primary Health Care Corporation and the Chairperson of the Tactical Community Command Group on COVID-19, as well as members of this committee, for providing support to the teams that worked on the field surveillance. We further thank Dr. Nahla Afifi, Director of Qatar Biobank (QBB), Ms. Tasneem Al-Hamad, Ms. Eiman Al-Khayat and the rest of the QBB team for their unwavering support in retrieving and analyzing samples and in compiling and generating databases for COVID-19 infection, as well as Dr. Asmaa Al-Thani, Chairperson of the Qatar Genome Programme Committee and Board Vice Chairperson of QBB, for her leadership of this effort. We also acknowledge the dedicated efforts of the Clinical Coding Team and the COVID-19 Mortality Review Team, both at Hamad Medical Corporation, and the Surveillance Team at the Ministry of Public Health.

## Author contributions

LJA conceived and designed the study, led the statistical analyses, and co-wrote the first draft of the article. HC contributed to study design, performed the data analyses, and co-wrote the first draft of the article. JAM led the viral genome sequencing analyses and AAA, YAM, and SY conducted these analyses. All authors contributed to data collection and acquisition, database development, discussion and interpretation of the results, and to the writing of the manuscript. All authors have read and approved the final manuscript.

## Data sharing

All relevant data are available within the manuscript and its supplementary materials.

## Supplementary Material

### Text S1. Details of the viral genome sequencing methods

Viral RNA was extracted using Quick-RNA Viral Kit (Zymo Research, Irvine, USA; Cat. No. R1041) and eluted in 30ul of nuclease-free water. RNA quality was assessed with real-time quantitative polymerase chain reaction (RT-qPCR) using SARS-CoV-2 (2019-nCoV) CDC qPCR Probe Assay Research Use Only (RUO) kit (Integrated DNA Technologies, USA; Cat number 10006713) and Luna Universal Probe One-Step RT-qPCR Kit (New England BioLabs, USA; Cat number E3006E) on Applied Biosystems 7500 Fast Real-Time PCR instrument (Applied Biosystems, CA, USA).

Next-generation sequencing (NGS) library construction was performed using CleanPlex SARS-CoV-2 Panel (Paragon Genomics, USA; SKU: 918012). Gel-size selection on a 3% agarose gel was utilized to prevent formation of adapter dimers. NGS libraries were quantified using KAPA Library Quantification Kit (Roche, USA; KK4824), and normalized, pooled, and sequenced on an Illumina MiSeq instrument using a paired-end 150bp kit (Illumina, USA; MS-102-2002). All procedures were implemented following manufacturers’ protocols.

Raw sequences were processed with CUTADAPT (v2.10) [1] to exclude the contaminating adapter sequences. Adapter trimming was performed using parameters -g CCTACACGACGCTCTTCCGATCT **-a** AGATCGGAAGAGCACACGTCTGAA **-A** AGATCGGAAGAGCGTCGTGTAGG **-G** TTCAGACGTGTGCTCTTCCGATCT **-e** 0.1 **-O** 9 **-m** 50 **-n** 2. Only paired reads with minimum length of 50bp were retained for analysis. The latter filtered reads were aligned to SARS-CoV-2 reference genome (NC_045512) using BWA-MEM [2]. FGBIO (v1.3.0) was subsequently used to remove PCR primer sequences from the resulting BAM file.

Variant calling and genotyping were performed with VarScan multi-sample mpileup [3] with the pileup file generated using SAMTOOLS mpileup (v1.10) [4] with --min-BQ 20 and --min-MQ 20 parameters. The mpileup2snp function of VarScan was then applied with the filtering parameters --min-var-freq 0.2, --min-coverage 5, and --min-avg-qual 20, to generate the final VCF file.

**Figure S1.**
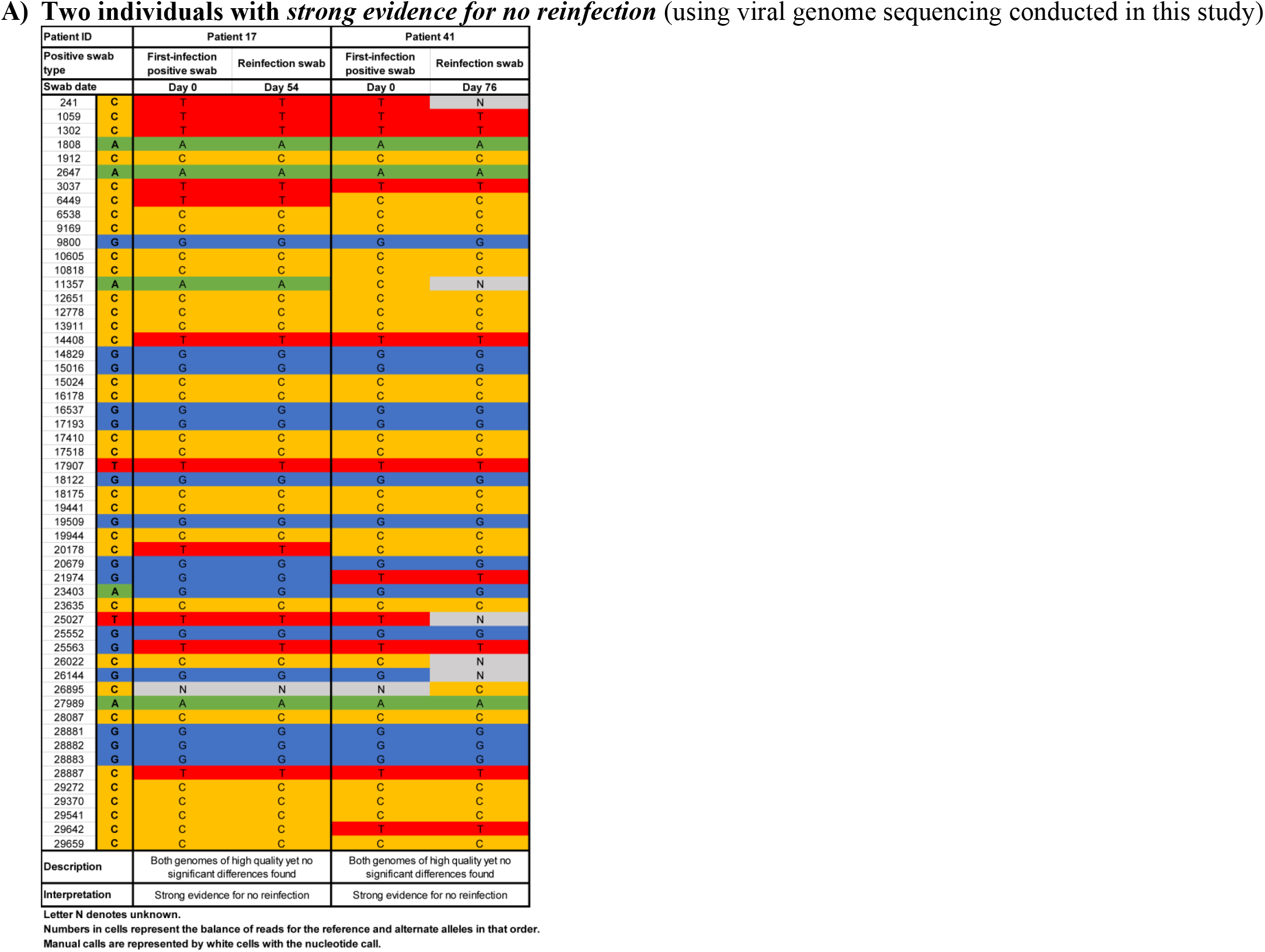
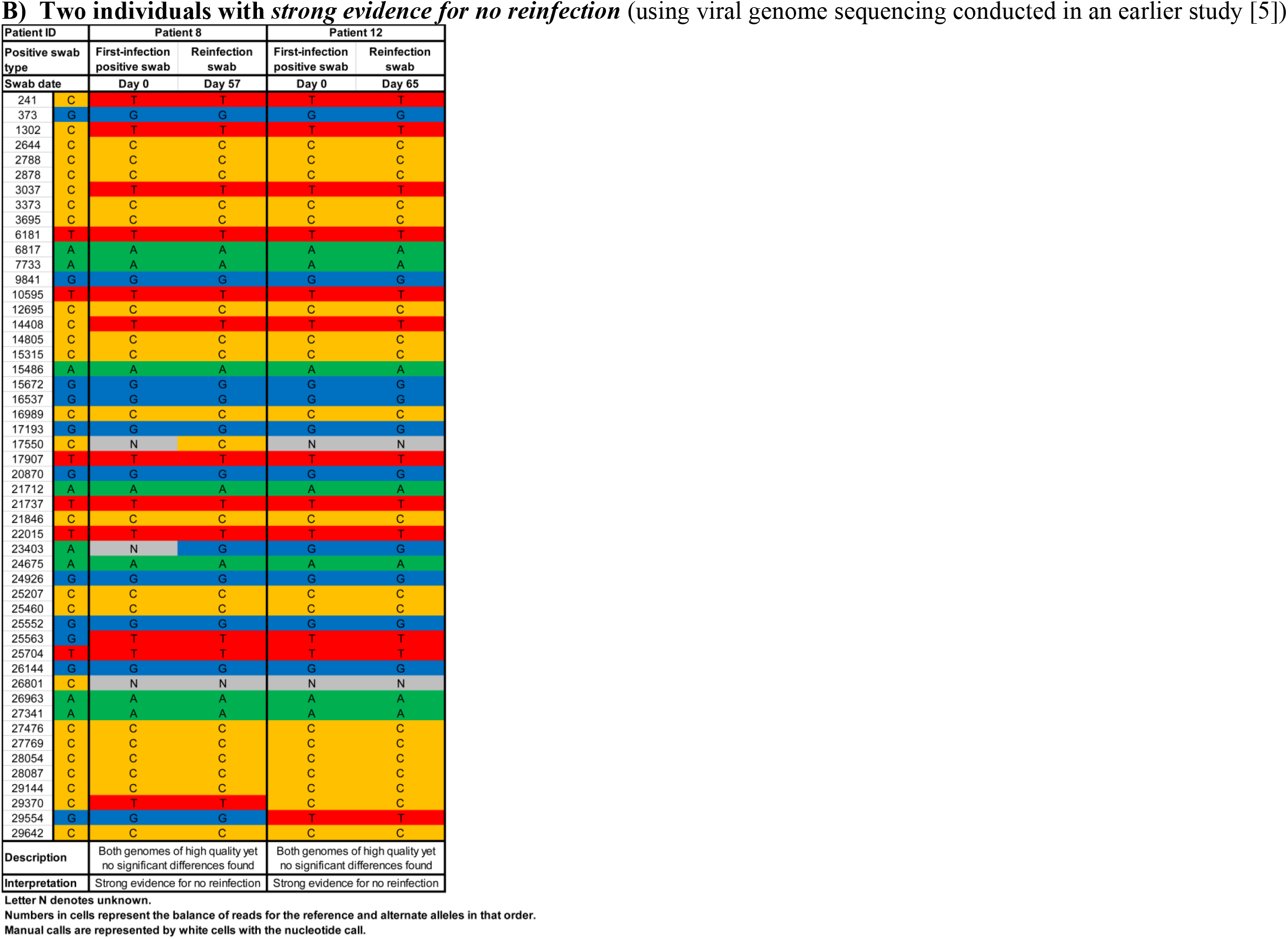
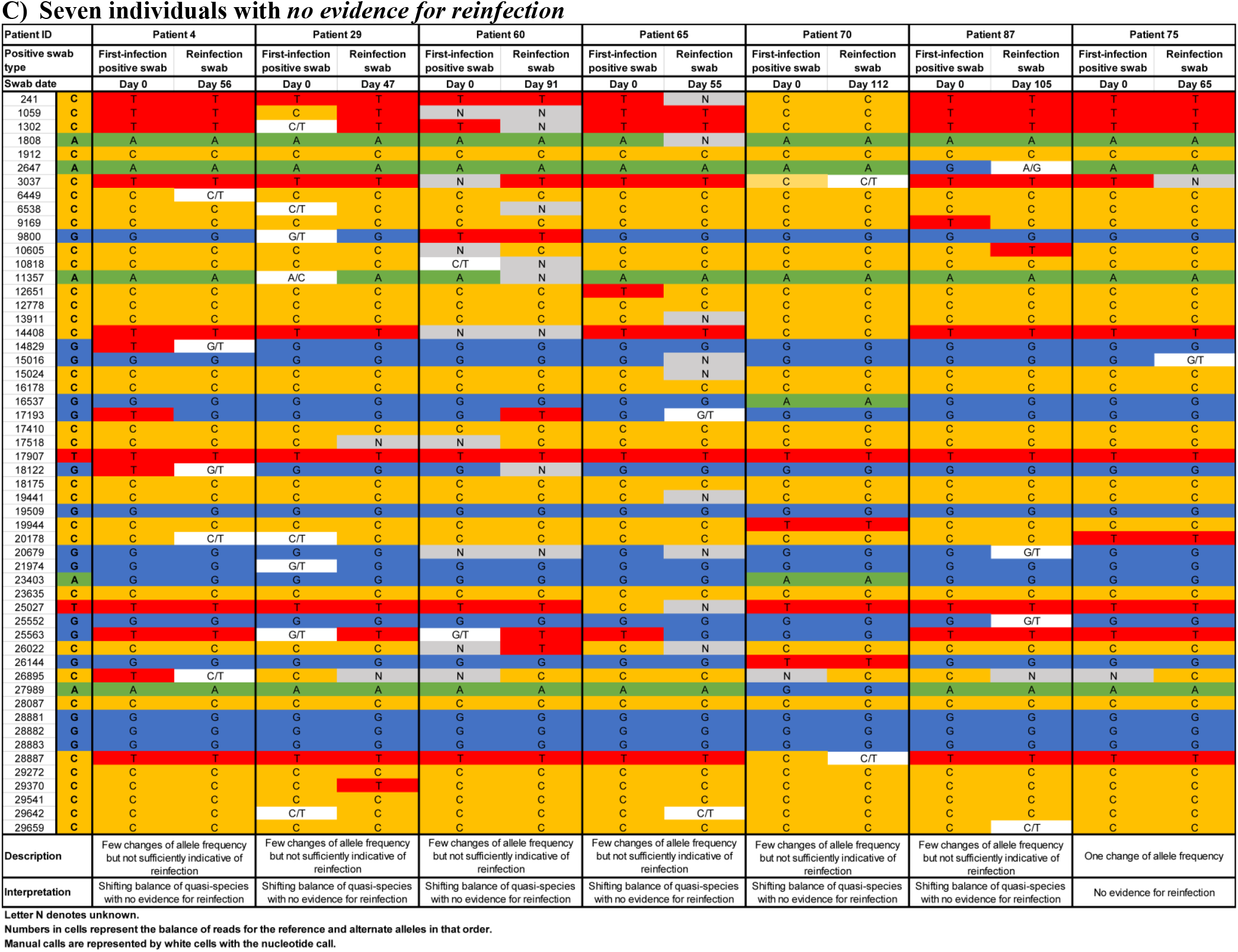
Viral genome sequencing analysis of the paired viral specimens of the primary-infection PCR-positive swab and the reinfection PCR-positive swab for the eleven cases with evidence not supporting occurrence of reinfection.

**Figure S2.**
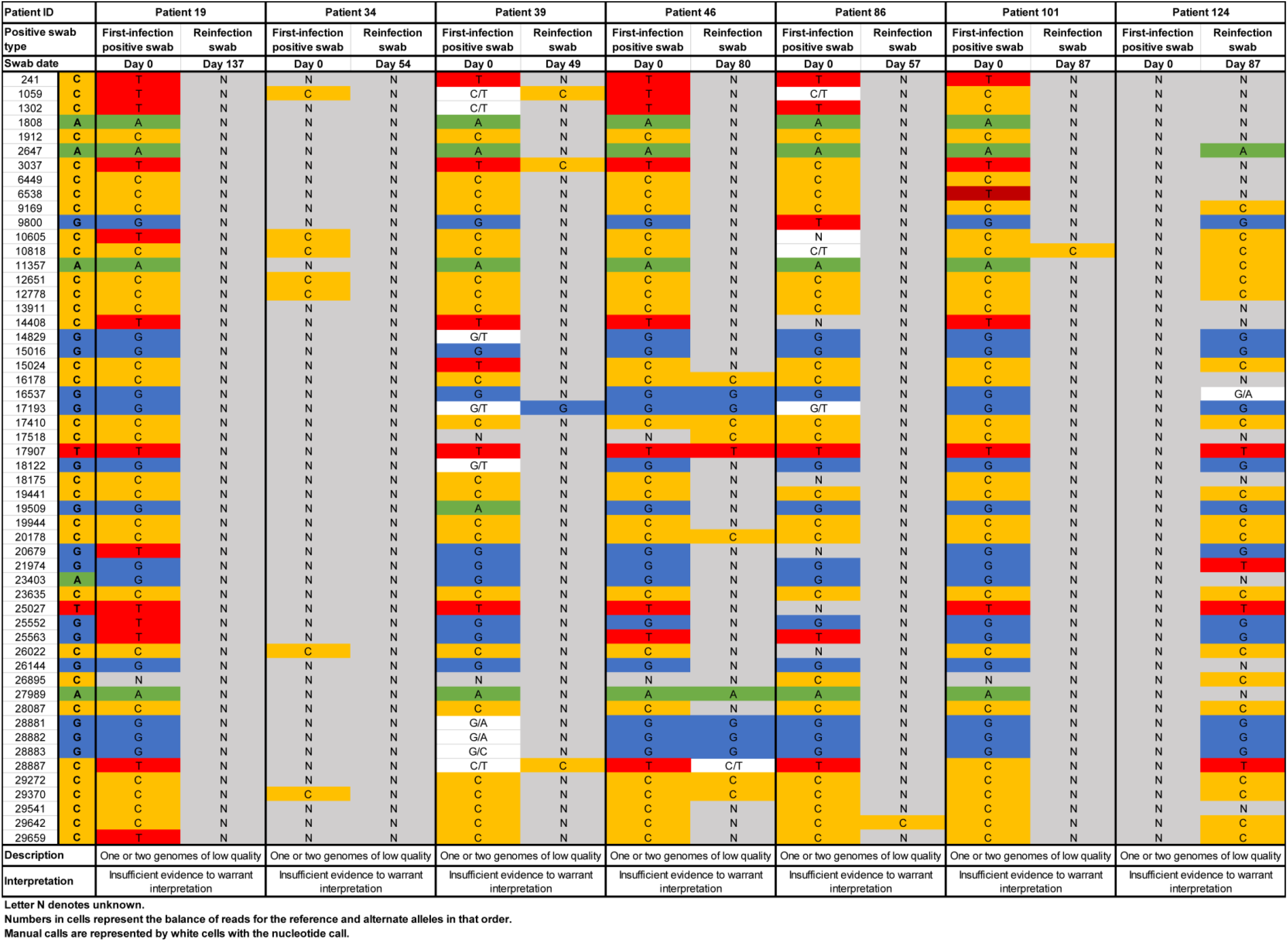
Viral genome sequencing analysis of the paired viral specimens of the primary-infection PCR-positive swab and the reinfection PCR-positive swab for the seven cases with insufficient genetic evidence to confirm the reinfection.

